# Functional evaluation of toothbrushes using two standardized models

**DOI:** 10.64898/2025.11.27.25341179

**Authors:** Mami Endoh, Yuki Tsuji, Tomoyo Jinushi, Shota Kurihara, Takuro Higuchi, Atsushi Takayanagi, Atsushi Yamagishi, Takato Nomoto

## Abstract

Manual toothbrushing is the most widely used method for daily plaque control. Although brushing techniques are crucial for plaque removal, it is very difficult to improve brushing techniques to maximize their effectiveness. Despite the variety of toothbrushes available, the objective evaluation of the functional index of a toothbrush remains elusive. This study aimed to develop a comprehensive and objective evaluation method for toothbrush function using standardizable models. For a comprehensive evaluation, we divided the evaluated surface into three elements (convex, concave, and flat). Further, we developed two standardized evaluation models: the smooth and interproximal surface model, which assesses the cleaning ability of both smooth and interproximal surfaces, and the **s**ingle half cylinder model (SHC model), which measures the following ability of toothbrush bristles on convex surfaces.

The results indicated that toothbrush G had equal or greater performance than toothbrush S across all tested angles in the smooth and interproximal surface model. The minimum following load necessary for effective cleaning without bristle bounce, was significantly lower for toothbrush G (1.25 N) than for toothbrush S (3.84 N), suggesting that toothbrush G is less dependent on brushing technique and more suitable for a wide range of individuals, regardless of their brushing skills.

Currently, few comprehensive and objective evaluation methods for toothbrush function have been reported using standardized evaluation methods. Moreover, a single evaluation model to assess toothbrush function might be inadequate due to varying oral cavity morphologies. However, the use of multiple models could potentially address this challenge. This study used two models to clearly demonstrate the performance differences between toothbrushes. This method could serve as a standard for the comprehensive evaluation of toothbrush functionality.

## 1 Introduction

Manual toothbrushes are the most widely used for daily plaque control. Notably, brushing time, frequency, and technique are crucial for effective toothbrushing. Although brushing techniques are crucial for plaque removal, it is very difficult to improve brushing techniques to maximize their effectiveness [1].

Despite the variety of toothbrushes used globally, no functional indices for objective evaluation of toothbrush performance exist. Previous methods for evaluating toothbrush function applied artificial dental plaque to a single type of dental model [2–4]. However, there are many kinds of dental models, and none of them have been standardized. Thus, objective evaluation of toothbrush function remains elusive, particularly because no specific dental models can account for the varying tooth and dental arch morphologies among individuals. The existence of various types of dental models also makes comparison with past results difficult. Although previous reports have used dental models for the evaluation of toothbrush functionality [2–4], comprehensive comparisons with these reports remain unclear since no unified evaluation criteria have been established. Moreover, no two models have identical tooth and dental arch morphologies.

For a standardized evaluation, we divided the evaluated surface into three elements (convex, concave, and flat). The single half cylinder model (SHC model) represents the convexity of the tooth surface [5–8], while the smooth and interproximal surface model represents the smooth and interproximal surfaces of the tooth [9–12]. These two models served as standardized evaluation methods for toothbrush functionality from multiple perspectives, such as the dental plaque removal ability of smooth and interproximal surfaces, and following ability on convex surfaces.

This study aimed to develop a comprehensive and objective evaluation method for toothbrush function using standardizable models.

## 2 Methods

### 2.1 Measuring model design

#### 2.1.1 Design of flat and concave surface

Evaluating the cleaning ability of toothbrushes on interproximal surfaces is essential because dental caries and periodontal disease occur mainly in these areas. However, evaluating the cleaning ability of a toothbrush on smooth surfaces is also important because the stain removal effect and oral comfort are primarily measured in these areas.

Therefore, we designed the smooth and interproximal surface model with an aluminum block. Two blocks of one long side of a flat aluminum block (64.0 mm width, 45.0 mm length, and 6.0 mm thickness) were processed as a circular sector with a radius of 4.0 mm and a center angle of 90°. They were fixed with their circular sector surfaces facing each other [12]. The circular sector radius was based on the average tooth width diameter of Japanese individuals [13].

#### 2.1.2 Design of convex surface

Given that there are convex surfaces on the teeth, it might be difficult for bristles to reach interproximal surfaces if the toothbrush does not move along those surfaces.

The movement of toothbrush bristles is dependent on the force applied when brushing the convex surfaces of the teeth [7, 8]. When the force is too low, the tips of the bristles bounce off when they contact the convex surface of teeth, which reduces cleaning effectiveness. Conversely, excessive force can cause gingival damage, as the bristles deflect to the point of losing their flexibility. This hardened part of the toothbrush might damage the oral cavity [8]. Given that the bristle tips do not move along the tooth surface properly when either excessive or low force is applied, an appropriate amount of force that allows the bristle to follow these surfaces properly is crucial for effective brushing. Consequently, we designed the single half cylinder model (SHC model) to determine the ideal force for a toothbrush to facilitate effective cleaning, based on the relationship between the applied force and the following ability. A stainless-steel semi-cylinder was placed on a flat stainless-steel plate [5–8]. The radius of the semi-cylinder was set at 4.0 mm, representing the average width of a Japanese individual’s teeth [13].

### 2.2 MEASURING DEVICE

Schematic view of the device is shown in **Fig 1**. Using a jig, the handle of the toothbrush was fixed to a hinge attached to the slider (FBW 2560XR+160L, THK, Tokyo, Japan).

**Fig 1.**
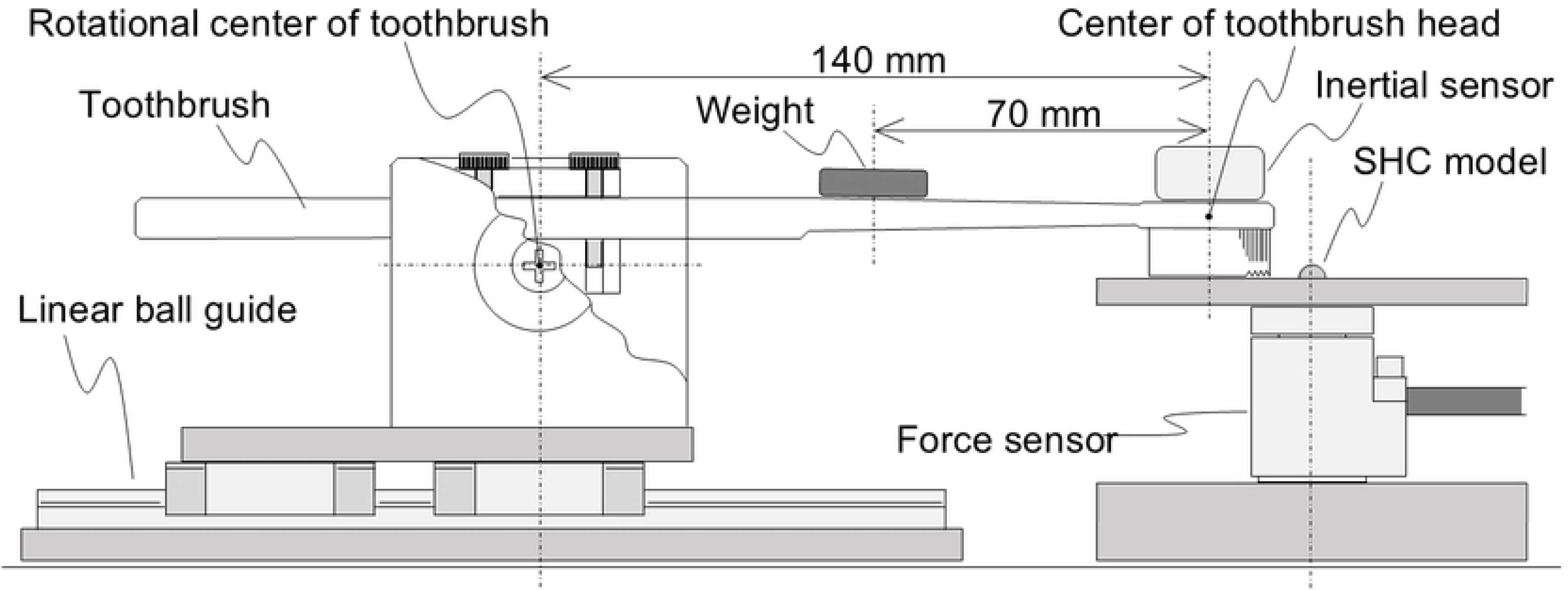
Schematic view of the experimental setup.

The distance from the hinge’s center (fulcrum) to that of the toothbrush head (point of load) was 140.0 mm. The weight was placed 70.0 mm from the center of the toothbrush head (point of effort), which was considered to be near the finger point of a toothbrush handle, close to the position where a person applies force during actual brushing.

### 2.3 Test toothbrushes

Two types of test toothbrushes were used: (S): toothbrush with 3 rows of flat bristles and (G): toothbrush with 4 rows of stepped bristles. **Fig 2** shows the appearance, and **Table 1** shows the specifications of each toothbrush.

**Fig 2.**
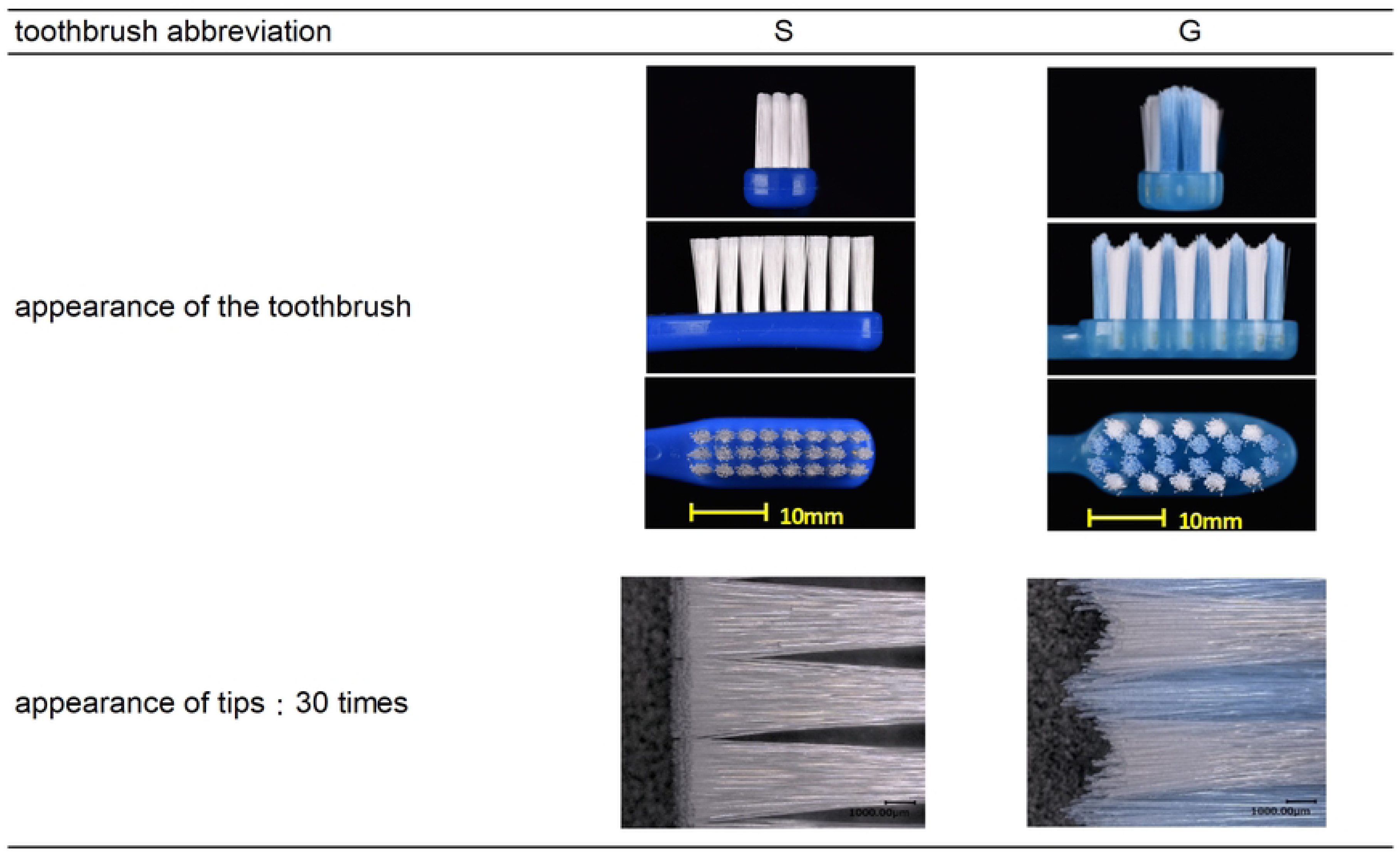
Appearance of toothbrushes.

**Table 1.**
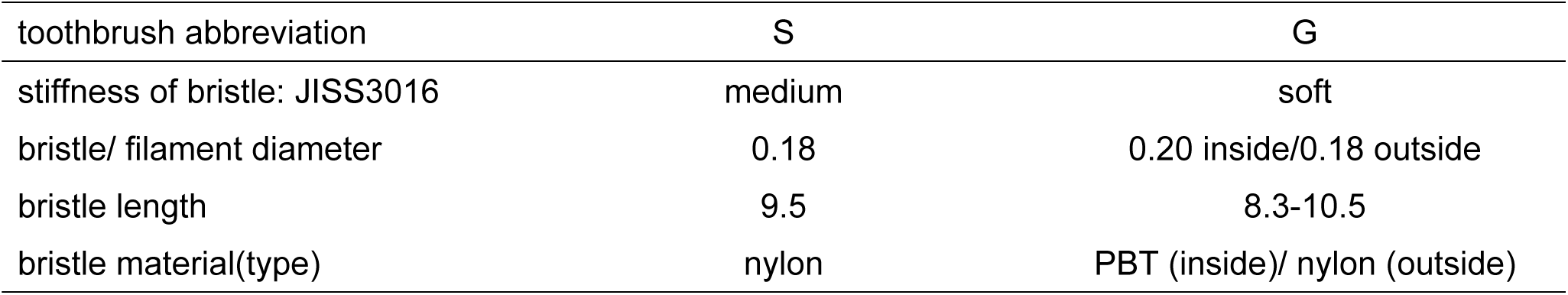
Specifications of toothbrushes.

| toothbrush abbreviation | S | G |
| --- | --- | --- |
| stiffness of bristle: JISS3016 | medium | soft |
| bristle/ filament diameter | 0.18 | 0.20 inside/0.18 outside |
| bristle length | 9.5 | 8.3-10.5 |
| bristle material(type) | nylon | PBT (inside)/ nylon (outside) |

### 2.4 Experiment with smooth and interproximal surface model

#### 2.4.1 Evaluation of cleaning ability

##### Measurement using the magnetic tape method

**Fig 3** shows the evaluation method of cleaning ability. We used the magnetic tape method [14] to evaluate the cleaning ability of the smooth and interproximal surfaces, which is determined by the amount of magnetic film that is peeled off by the bristle tips during cleaning. When the toothbrush, the smooth and interproximal surface model was placed in the evaluation device, magnetic tape was affixed around the curved surface from the smooth surface of the model, where the bristle tips were expected to make contact. The magnetic tapes were attached with double-sided tape, with the magnetic film on top, to prevent overlapping and gaps.

**Fig 3.**
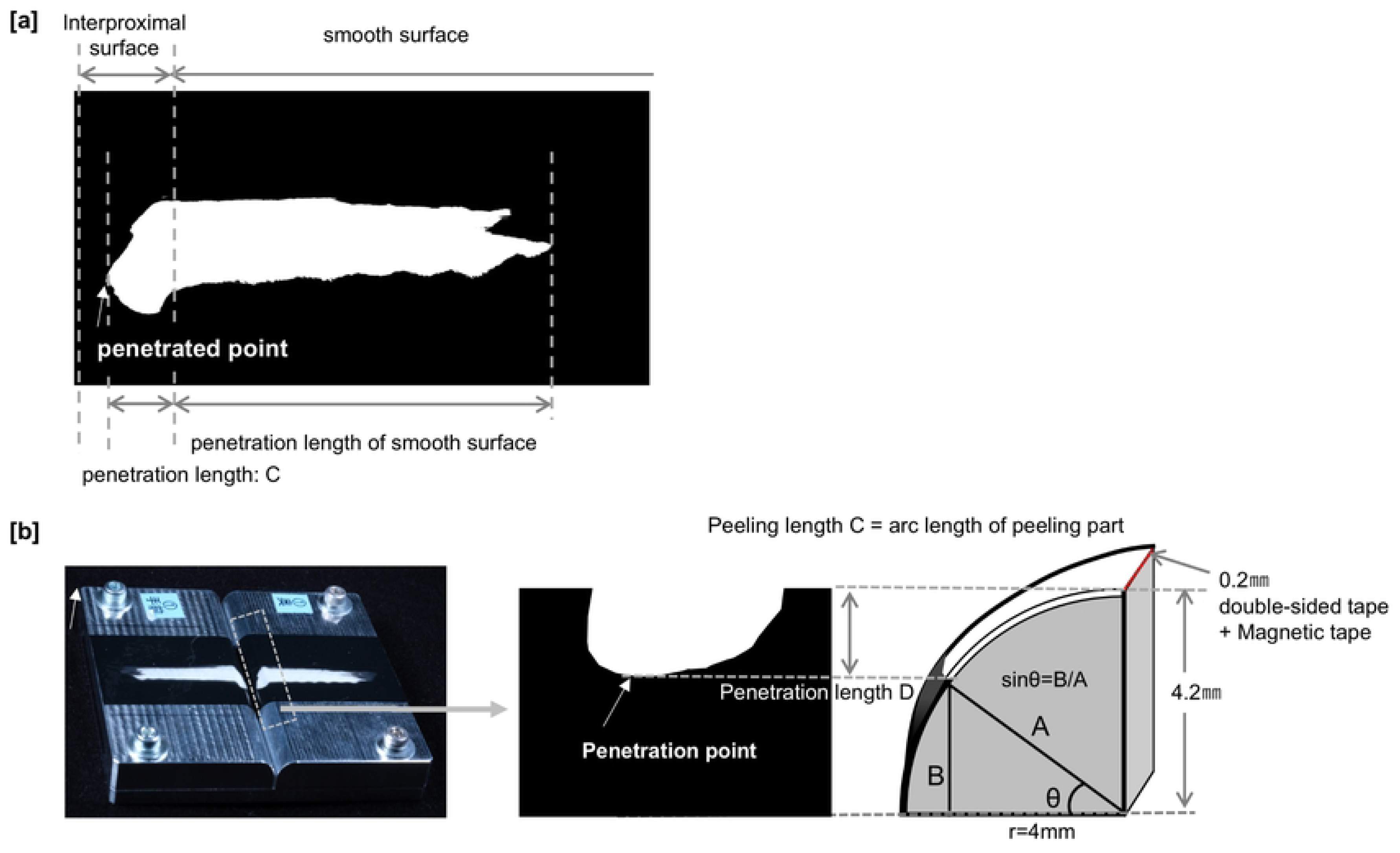
Calculating method of penetration length in interproximal surface. (a) magnetic tape affixed to an acrylic flat plate after removal from the model. (b) Penetration length of magnetic tape in the interproximal surface when attaching the model (schematic diagram).

##### Brushing stroke, frequency, and force

The measurements were conducted with a brushing stroke of 40.0 mm and a brushing frequency of 2 Hz per unit time, based on previous studies [6–12]. The force during the experiment with the smooth and interproximal surface model was 2 N, based on the average force during actual brushing [15, 16]. The weight was adjusted to a set value while monitoring the force applied to the toothbrush during actual brushing using a force sensor (030YA151U, Leptrino, Nagano, Japan) placed at the bottom of the smooth and interproximal surface model. Notably, we used 1.0 g of toothpaste Guard Halo ^®^ (Kao Corporation, Tokyo, Japan)—which is commonly available in Japan—in this study. The measurements were conducted three times, comprising 200 strokes/100 s.

The model was tilted 0°, 15°, and 30° to the brush sweep axis and photographed at each angle. The angle of contact between the model and toothbrush was photographed using a digital camera (OM-D E-M5 Mark Ⅲ, OM Digital Solutions, Tokyo, Japan) with a microlens (Micro NIKKOR 60 mm f/2.8D, NIKON, Tokyo, Japan). The camera was set up such that its imaging plane was perpendicular to that of the model and the brushing direction. The toothbrush contact perpendicular to the model surface was defined as 0°.

#### 2.4.2 Measurement of peeled area and penetration length

The model was removed from the measuring device following completion of the test and subsequently rinsed with water to remove the dentifrice. The magnetic tape affixed to a white acrylic flat plate was photographed using a digital camera (D5600, NIKON, Tokyo, Japan) with a microlens (AF-S DX Micro NIKKOR 40 mm f/2.8 G, Nikon, Tokyo, Japan). After converting the resulting image to grayscale, it was analyzed using ImageJ^®^ (NIH, USA). The analysis was conducted independently for the smooth and interproximal surfaces. After measuring each peeled area of the magnetic tape, the penetration length was calculated using the method described in a previous report [12]. Given that the total thickness of the magnetic tape and the double-sided tape was 0.2 mm, the radius of the circular sector was set to a total of 4.2 mm with a radius of 4.0 mm at the interproximal surface of the model when performing the calculations (**Fig 3)**. Further, t-tests and Bonferroni corrections were performed for the penetration length and peeled area with respect to toothbrush type and head angle. A t-test was used to test for between-group differences, while a Bonferroni correction was used when testing for differences between three groups.

### 2.5 Experiment with the single half cylinder model

#### 2.5.1 Evaluation of the following load

For measurement of toothbrush movement, a 6-axis inertial sensor (IMU; AMWS020^®^, ATR-Promotions, Kyoto, Japan) was attached to the toothbrush head. The measurement axes were set as follows: the Y-axis for the front-back direction (stroke direction), the X-axis for the left-right direction, and the Z-axis for the up-down direction. Measurements were conducted with a specialized software (Sensor Controller^®,^ ATR-Promotions, Kyoto, Japan) at a sampling rate of 1,000 Hz. The load for the toothbrush was set to 0.5–7 N, based on a previous study [5]. The weight was adjusted to a specific value while monitoring the force applied to the toothbrush during actual brushing using a force sensor (030YA151U, Leptrino, Nagano, Japan) placed at the bottom of the model, same as the smooth and interproximal surface model. The brushing time was set to approximately 20 s. Furthermore, the measurements were conducted with a brushing stroke of 40.0 mm and a brushing frequency of 2 Hz per unit time, based on previous studies [6–12]. Each measurement was conducted three times. We performed t-tests to analyze the penetration length and peeled area with respect to the toothbrush type and head angle.

#### 2.5.2 Evaluation of the following ability of the toothbrush head

The analysis was performed for 10 strokes/5 s of brushing. The velocity of the stroke direction was calculated from the Y-axis acceleration and used as the basis for measuring the head motion. For the vertical motion of the toothbrush head, displacement was obtained by second-order integration of the Z-axis acceleration, which is the vertical acceleration during one stroke. The maximum value of the toothbrush head movement in the positive direction and the minimum value in the negative direction were extracted, and the sum of the absolute values of the maximum and minimum values during one round trip was used as the head amplitude [9] to evaluate the head behavior. The maximum and minimum values were obtained during one stroke, and the average value of 10 strokes was calculated.

#### 2.5.3 Definition and calculation of the following load

The force required for the toothbrush to achieve an appropriate cleaning effect was defined based on the relationship between the force and the followability of the SHC model. The minimum effective force was determined when the amplitude of the radius of the SHC model was <4 mm. The minimum effective force was calculated using the linear function slope between the measured values of the force before and after a toothbrush amplitude of 4 mm.

#### 2.5.4 Observation of the toothbrush head and bristle tips during brushing

Whilst measuring, video recordings were made for each toothbrush during brushing, using a digital camera (Z6, Nikon, Tokyo, Japan) with a lens (AF-S Micro NIKKOR 60 mm f/2.8 G ED, Nikon, Tokyo, Japan). Shooting conditions were as follows: frame rate: 120 fps, shutter speed: 1/4,000 s, and f8.

## 3 Results

### 3.1 SMOOTH AND INTERPROXIMAL SURFACE MODEL

#### 3.1.1 Peeled profile and contact area in each angle

**Fig 4** shows the peeled profile of the magnetic tape, which was the result of the experiment at 0° using the smooth and interproximal surface model.

**Fig 4.**
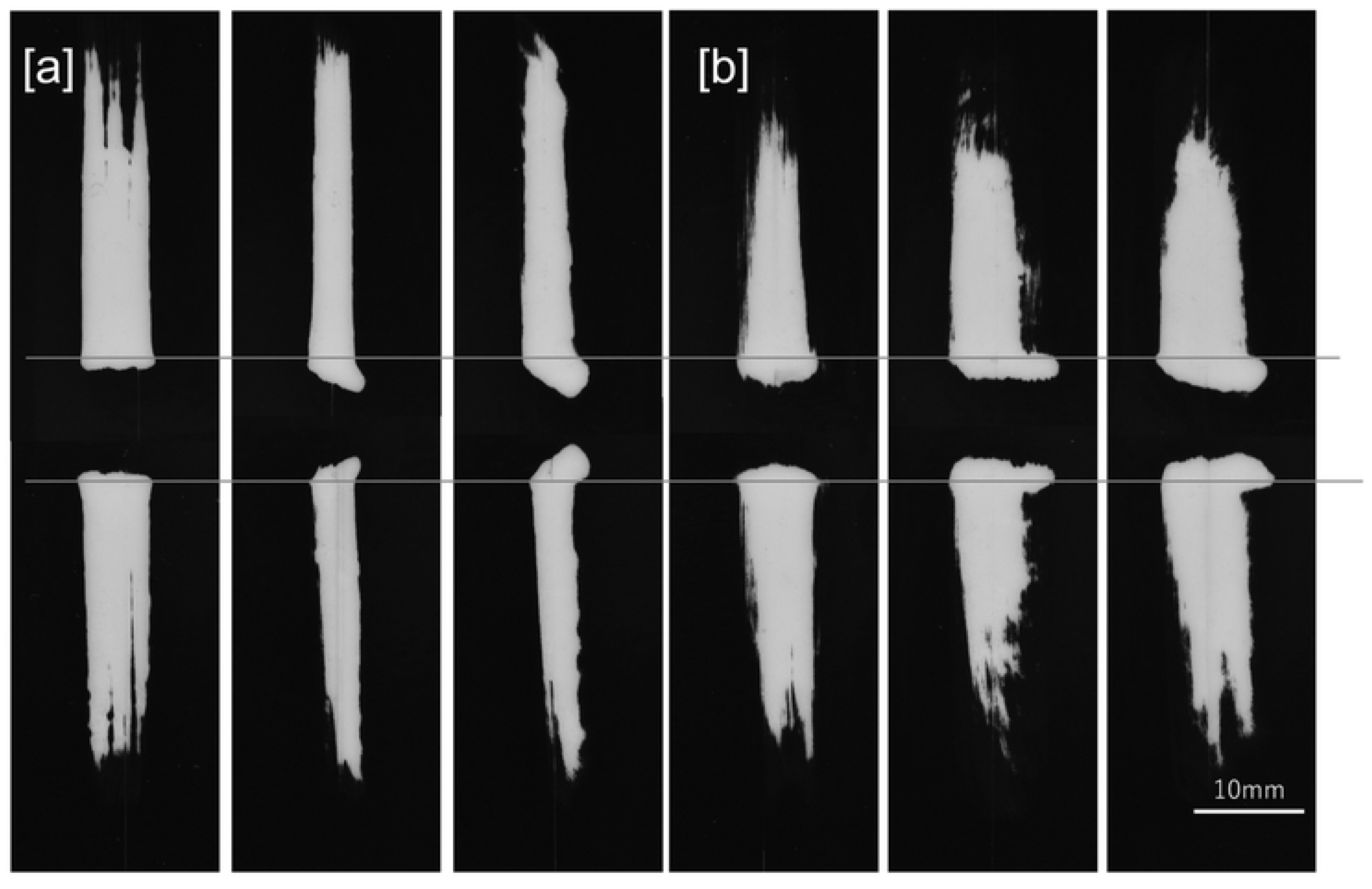
Peeled profile image of magnetic tape after experiment using the smooth and interproximal surface model. [a] toothbrush S, and [b] toothbrush G.

The contact between the bristle tips of the toothbrush and the smooth surface model is shown in **Fig 5**. Toothbrush S had the largest contact area at a head angle of 0°. At 0°, there was minimal deflection of the bristle tips; however, as the head angle increased, deflection was observed on the side of the toothbrush. For toothbrush G, there was minimal deflection of the bristle tips at 0° and 15°. Moreover, deflection was observed on the side and center of the toothbrush at 30°.

**Fig 5.**
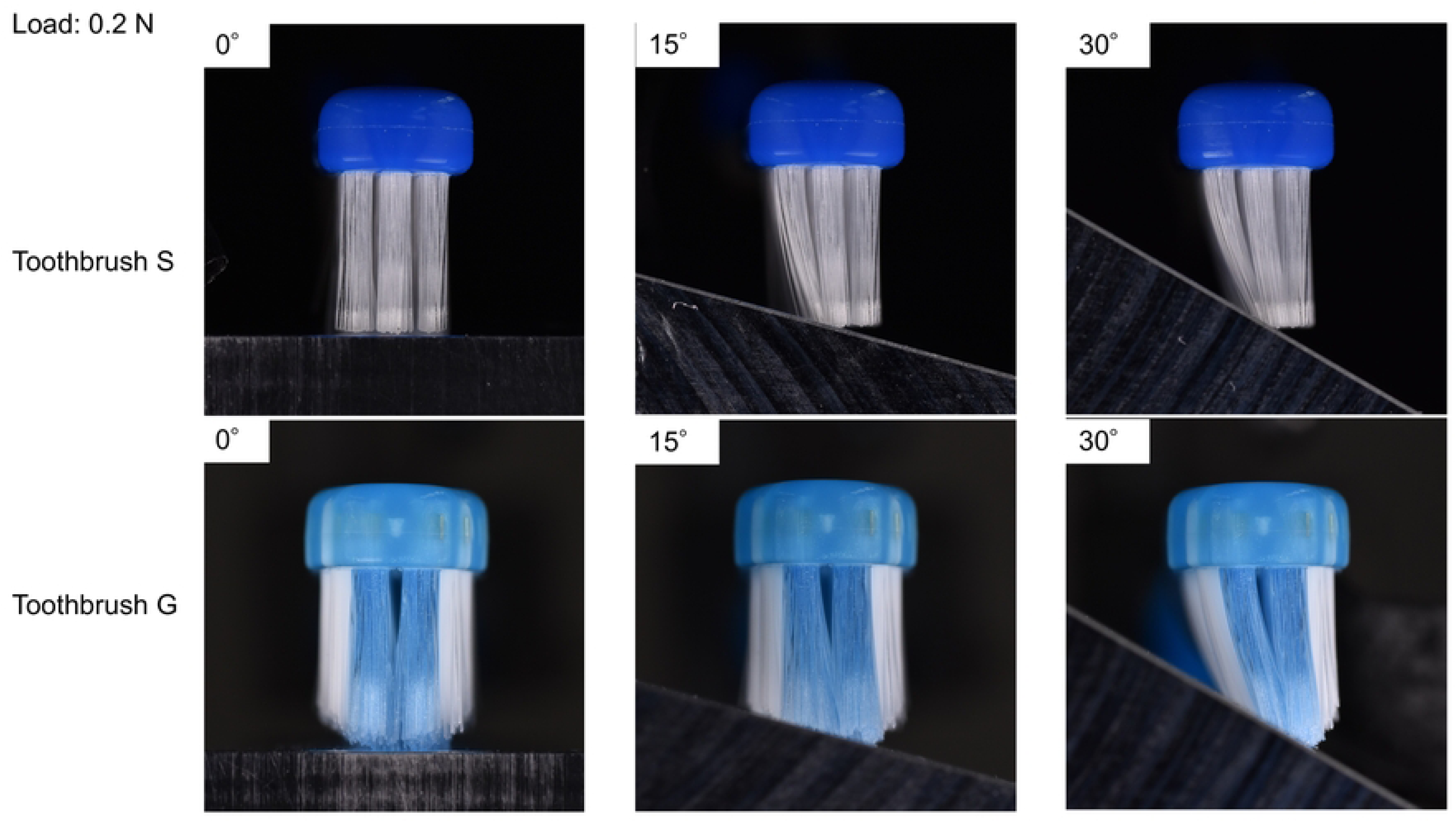
Contact state of the toothbrushes with the model.

#### 3.1.2 Analysis of smooth surface model

**Fig 6** shows each peeled area of the smooth surface model of toothbrushes S and G. Toothbrush S had the largest peeled area (253.9 mm^2^) at 0°, with significant differences between angles 0° and 15° (p<0.01), and also between angles 0° and 30° (p<0.05). Toothbrush G had the largest peeled area (310.5 mm^2^) at 30°, with significant differences between angles 0° and 30°, and also between angles 15° and 30° (p<0.05).

**Fig 6.**
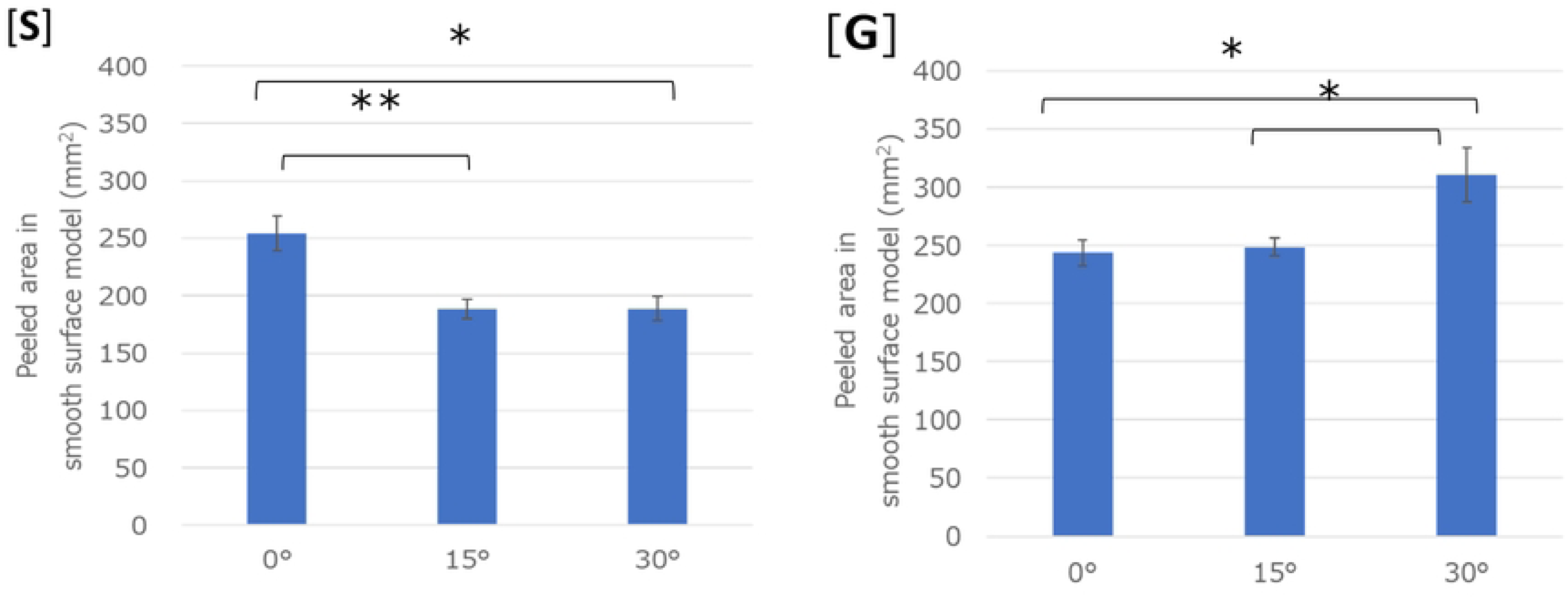
Peeled area in smooth surface model at each angle. [S]: toothbrush S and [G]: toothbrush G (**p<0.01, *p<0.05).

Toothbrush G had a significantly larger peeled area than toothbrush S at angles 15° and 30° (p<0.01) (**Table 2**). Overall, toothbrush G showed equal or greater peeled area than that of toothbrush S in all angles.

**Table 2.**
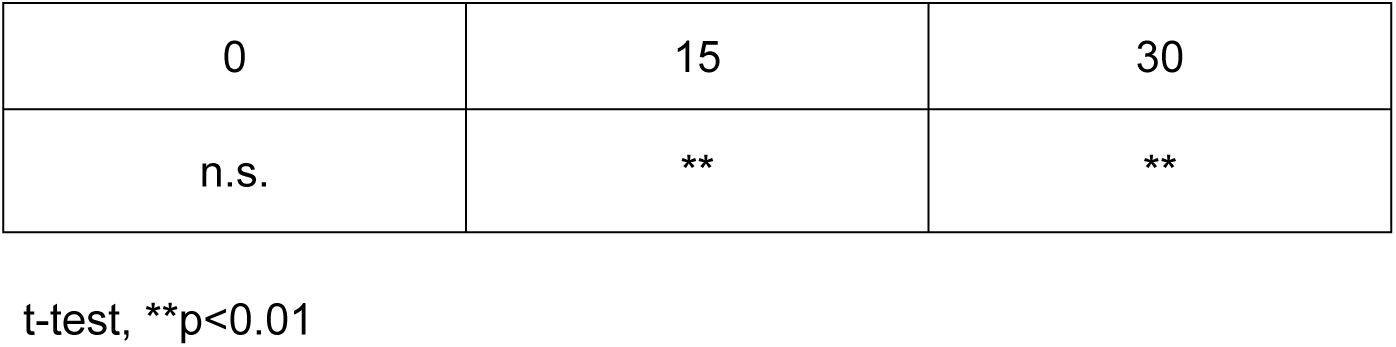
Peeled area in smooth surface model comparison of toothbrushes S and G at each angle.

#### 3.1.3 Analysis of the interproximal surface model

**Fig 7** shows the peeled areas of the interproximal surface model of toothbrushes S and G. The peeled areas of both toothbrushes significantly increased with increasing angles. The largest peeled area of toothbrush S (13.7 mm^2^) and G (25.3 mm^2^) was observed at 30°. For toothbrush S, significant differences were observed between angles 0° and 15°, 0° and 30°, and 15° and 30°(p<0.01). However, for toothbrush G, significant differences were observed between angles 0° and 15° (p<0.05), 0° and 30° (p<0.01), and 15° and 30° (p<0.01).

**Fig 7.**
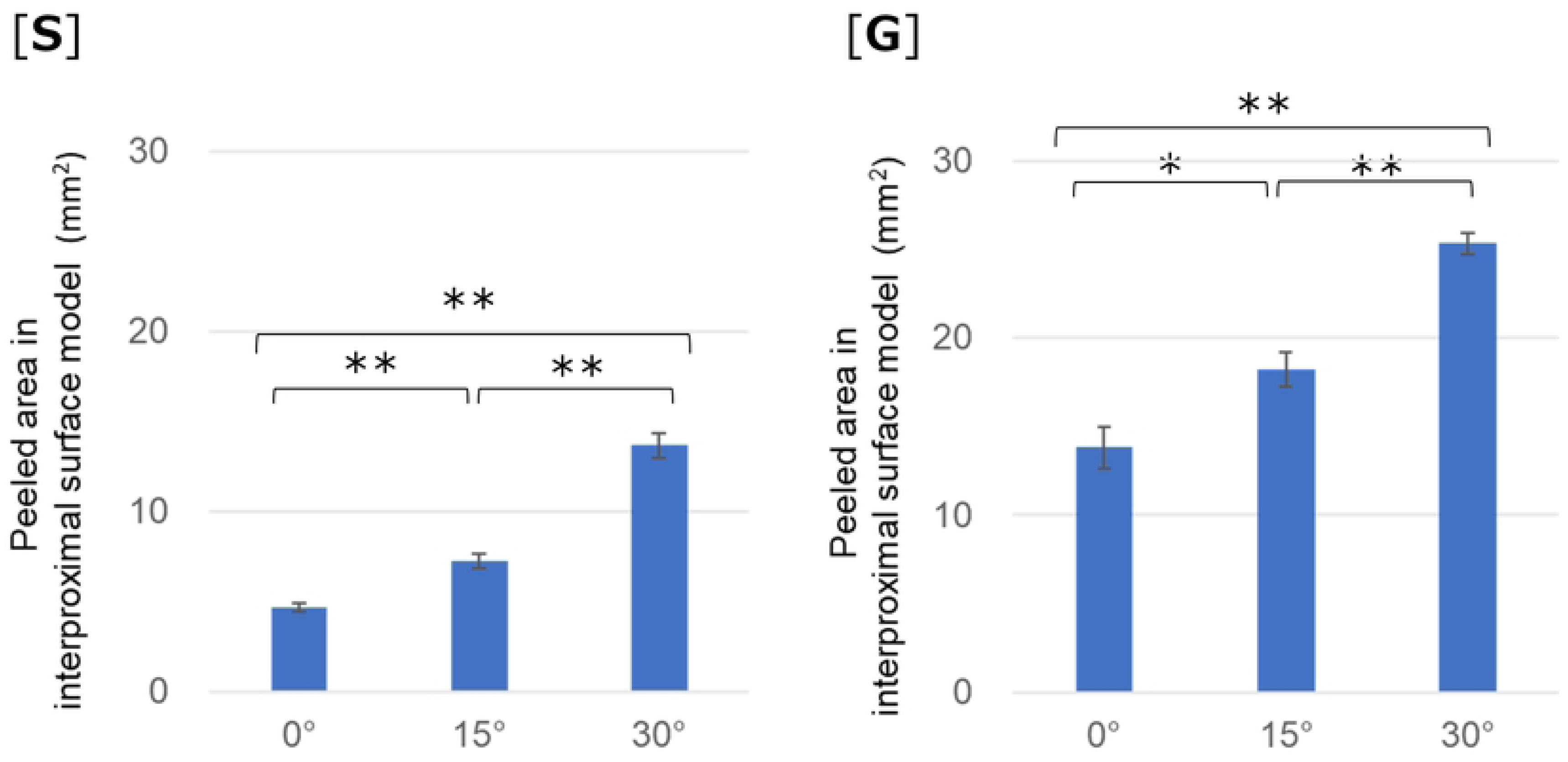
Peeled area in the interproximal surface model at each angle. [S]: toothbrush S and [G]: toothbrush G (**p<0.01, *p<0.05).

Toothbrush G had a significantly larger peeled area than toothbrush S at all angles (p<0.01). (**Table 3**) Overall, toothbrush G showed a greater peeled area than that of toothbrush S at all angles.

**Table 3.**
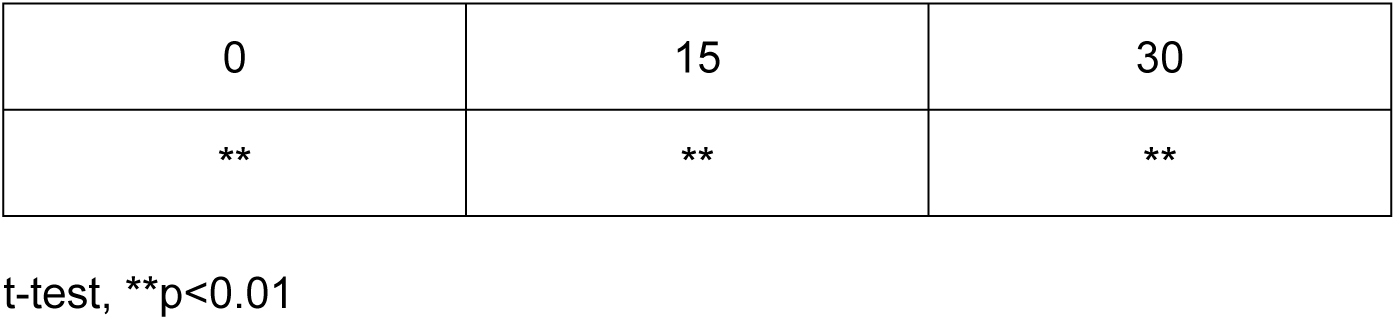
Peeled area in interproximal surface model comparison of toothbrushes G and S at each angle.

**Fig 8** shows the penetration lengths of toothbrushes S and G. The penetration length increased significantly with increasing toothbrush head angle, with significant differences between 0° and 15°, 0° and 30°, and 15° and 30° at toothbrush S (p<0.01). For toothbrush G, significant differences were observed between angles 0° and 30°, and also between angles 15° and 30° (p<0.01). Significant difference was observed between toothbrushes S and G at 0° (p<0.01) **(Table 4).**

**Fig 8.**
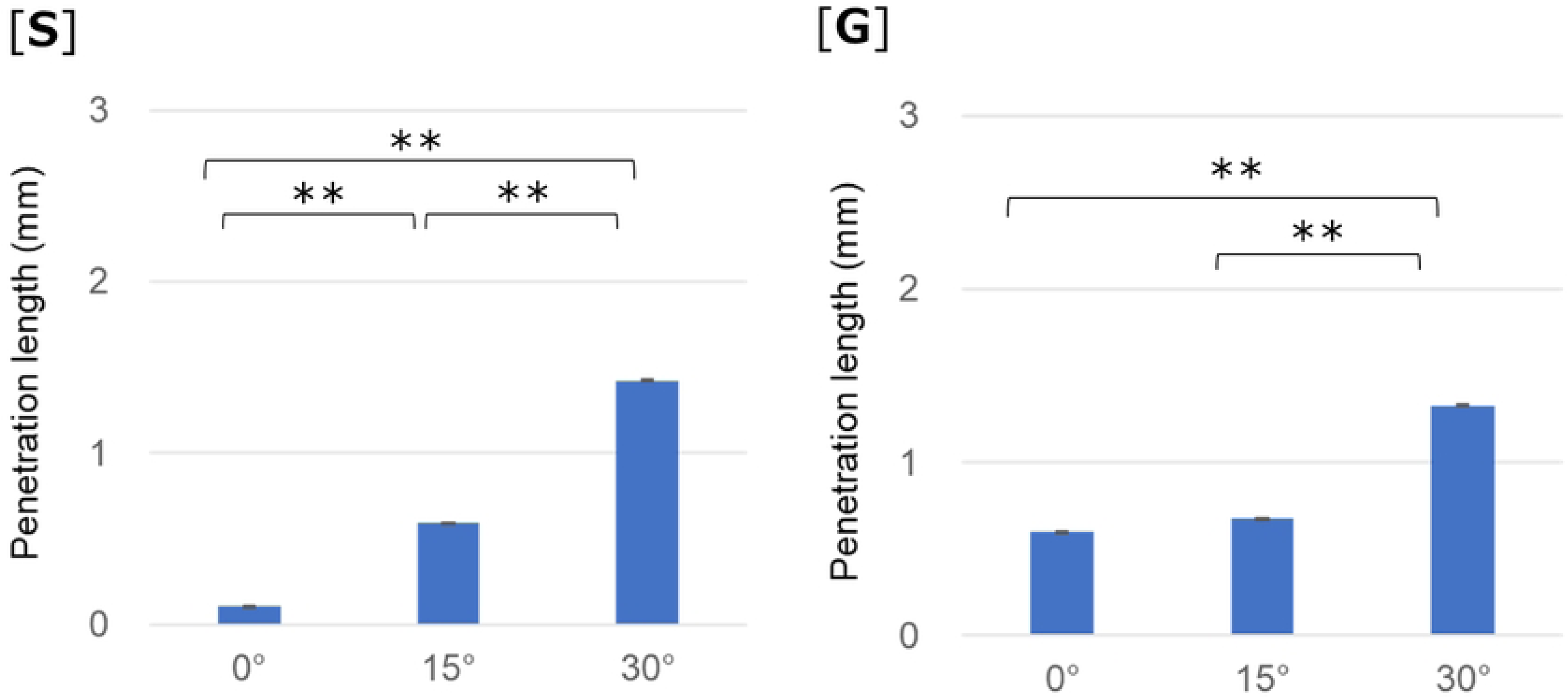
Penetration length in the interproximal surface model at each angle. [S]: toothbrush S and [G]: toothbrush G (**p<0.01, *p<0.05)

**Table 4.**
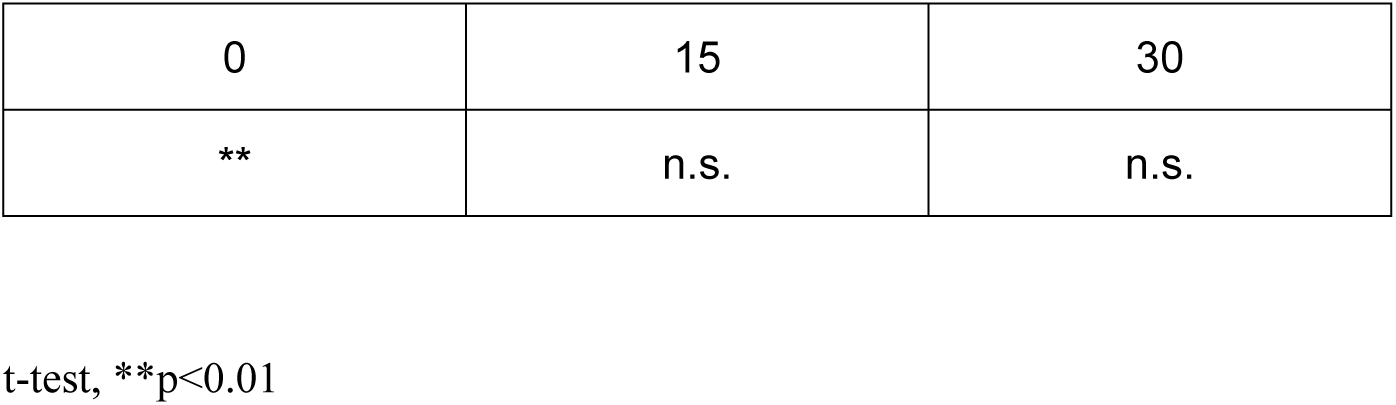
Penetration length in interproximal surface model comparison of toothbrushes G and S at each angle.

### 3.2 LOAD AND FOLLOWING ABILITY

**Fig 9** shows the relationship between the load and amplitude of toothbrushes S and G. **Fig 10** shows the images of the toothbrush making contact with the SHC model. The minimum load required to achieve appropriate cleaning effect was estimated to be 3.84 N (SD 0.45 N) for toothbrush S and 1.25 N (SD 0.10 N) for toothbrush G, as head amplitude was <4 mm, which is the radius of the semicylinder of the SHC model at that force. Toothbrush G requires a lighter load than toothbrush S. Each toothbrush required a load of approximately 7 N. Furthermore, toothbrush G had a significantly lower minimum load (p < 0.05) than toothbrush S did. Whenever the load was below the minimum load required, the bristle tips of the toothbrush bounced off as it passed through the SHC model and did not make contact with the surface, resulting in large amplitude values. As shown in **Fig 10**, both bristle tips of the toothbrush bounced off when it reached the model surface at a load of 0.5 N. Conversely, toothbrush bristles were flexed and compressed without bouncing off at 4 N. This deflection increased when the load was further increased to 7 N (**Fig 10**).

**Fig 9.**
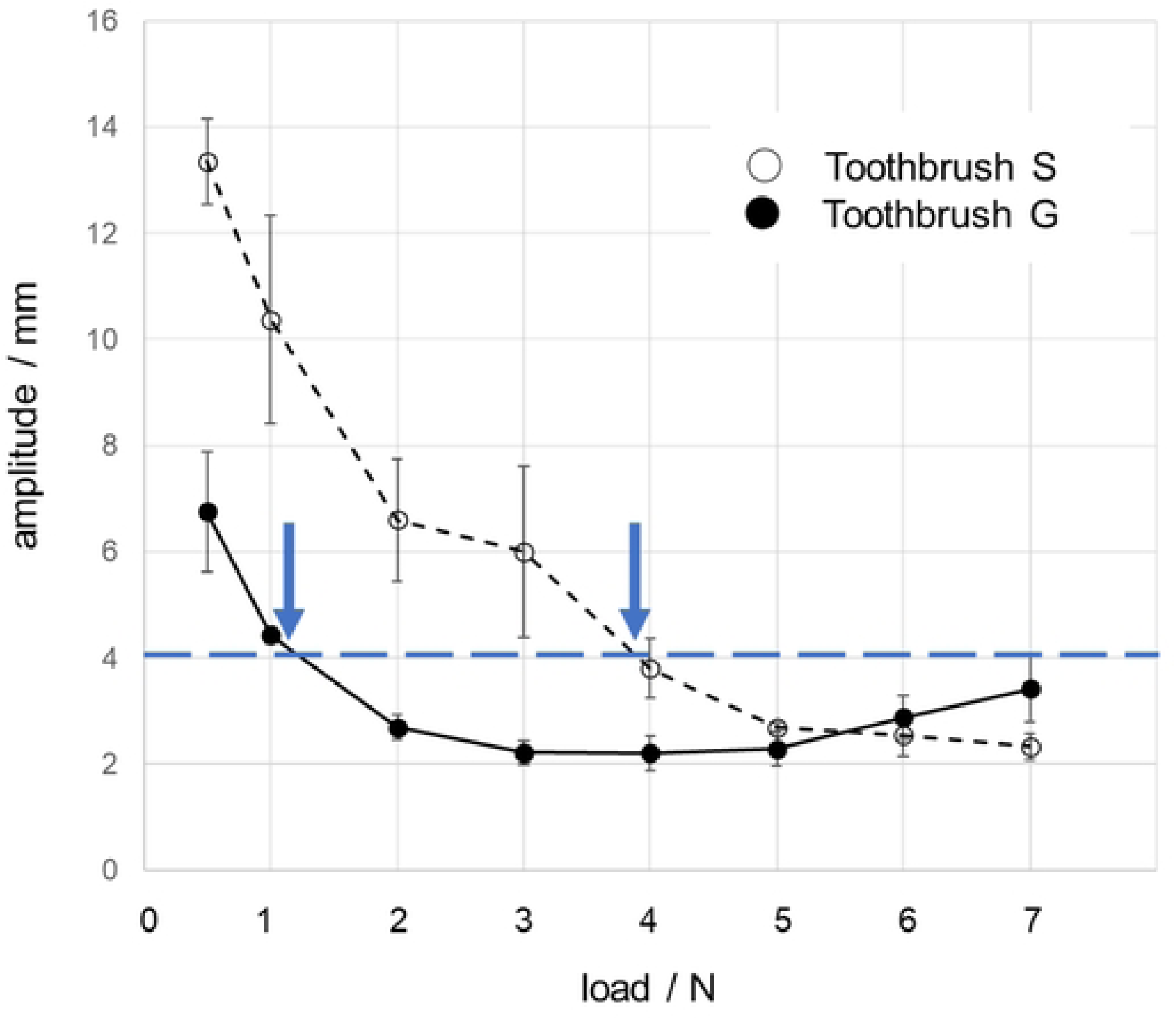
Relation between load and amplitude. ○ : toothbrush S, ●: toothbrush G, ↓: minimum following load of each toothbrush

**Fig 10.** Change in toothbrush behavior at each load during the experiment using the SHC model.

## 4 Discussion

Notably, an effective toothbrush should be able to brush efficiently, reach the interproximal surface, and move along uneven tooth surfaces without the bristles bouncing off. Furthermore, some people do not orient the angle of the toothbrush head to fit the tooth surface when brushing, while others do[1]. Consequently, evaluating the functionality of the toothbrush when the angle of the toothbrush head is changed is essential. Therefore, we evaluated the functionality of the toothbrushes using multiple models for a comprehensive assessment.

Toothbrush S had the largest peeled area (253.9 mm^2^) on the smooth surface model at 0°. Significant differences were observed between angles 0° and 15° (p < 0.01), and between 0° and 30° (p < 0.05). For toothbrush S, the decrease in the peeled area at 15° and 30° compared to 0°, suggests a smaller contact area between the toothbrush and model. Toothbrush G showed the largest peeled area (310.5 mm^2^) at 30°, and a significant difference was observed between angles 0° and 30°, and also between angles 15° and 30° (p < 0.05). As shown in **Fig 5**, toothbrush G had short, dome-shaped bristles on both sides. Minimal deflection was observed on the sides of toothbrush G when it made contact with the smooth surface model at angles 0° and 15°. However, no difference was observed in the peeled areas between angles 0° and 15°. Conversely, the peeled area increased at 30° compared to that at 0° and 15°. This increase was possibly caused by deflection of the outer bristles.

Toothbrush G showed a significantly larger peeled area than toothbrush S at 15 ° and 30° (p < 0.01) in the smooth surface model, indicating that toothbrush G has a higher cleaning ability than toothbrush S on a smooth surface.

In the interproximal surface model, the peeled area of both toothbrushes increased significantly with the head angle. The largest peeled area for both toothbrushes in the interproximal surface model was observed at 30°.

Overall, toothbrush G had a significantly larger peeled area than toothbrush S at all angles (p < 0.01). Although the outer bristles on both sides of toothbrush G often limit the tips’ ability to reach the interproximal surface, it could penetrate the interproximal surface because of its shorter bristles on both sides. Thus, toothbrush G showed a larger peeled area than toothbrush S.

The penetration length increased significantly with increasing head angle for toothbrush S. Significant differences were observed between angles 0° and 30°, and between angles 15° and 30° in the penetration length for toothbrush G (p < 0.01). Bristle deflection on both sides of toothbrush S increased with increasing angle, suggesting that the penetration length of toothbrush S increased with deflection of the bristles. For toothbrush S, a shorter penetration length was observed at 0°, suggesting a minimal deflection of the bristles. In addition, minimal deflection was observed on the side bristles of toothbrush G when it made contact with the model at angles 0° and 15°. Overall, no significant differences were observed between the penetration lengths at angles 0° and 15°. Penetration length increased at 30° because there was deflection in the bristles on both sides. Our results are similar to those obtained in an experiment using the smooth surface model. No significant differences in the penetration lengths between angles 0° and 15° of toothbrush G were observed, whereas a significant difference was observed in the peeled area in interproximal surface between 0° and 15° of toothbrush G. This finding might be due to the differences in the number of bristle tips that penetrate interproximal surfaces. Moreover, we expected that the number of bristle tips that can penetrate should increase with increasing head angle, and that a higher number of bristle tips would lead to a larger peeled area.

The penetration length of toothbrush G was larger than that of toothbrush S, with a significant difference observed at 0° (p < 0.01). Deflecting all bristles, except for the penetrating bristle, is essential to achieve a longer penetration length. Toothbrush G had short bristles on both sides, which allowed more bristles to penetrate the interproximal surface. Therefore, toothbrush G showed longer penetration length than toothbrush S.

At 0°, toothbrush S demonstrated a small peeled area and a short penetration length at the interproximal surface, whereas toothbrush G had a large peeled area and a long penetration length. We recommend toothbrush G for people who have poor brushing techniques because of its large cleaning areas and ability to penetrate the interproximal surface.

To remove the plaque effectively, the bristles of a toothbrush must be able to properly follow the curved surfaces of the teeth. However, effective use of a toothbrush is challenging when the force is less than the minimum required because the toothbrush head bounces off, and the bristle tips do not make contact with the surface of the SHC model. Therefore, toothbrushes with a high minimum force requirement necessitate a strong brushing force for the bristle tips to move along the curved tooth surfaces.

The minimum load for effective bristle contact in toothbrush S was 3.84 N, and that of toothbrush G was 1.24 N. The minimum load required in toothbrush G was significantly lower than that for toothbrush S. This result indicates that toothbrush G has a wider range than toothbrush S. Thus, a toothbrush with a low minimum force requirement and a wide range will be less dependent on the brushing force applied, facilitating effective brushing.

Effective brushing requires controlling the force and angle of the toothbrush on the tooth surface. Given that not everyone can use a toothbrush efficiently, the design of a toothbrush is crucial. Toothbrush G showed equal or better performance than toothbrush S across all experimental conditions, indicating that toothbrush G could be beneficial for people who are faced with the challenge of controlling the brushing force and angle to the tooth surface. However, toothbrush S is considered suitable for people who can effectively control their brushing force and angle because it is effective for optimal performance under those specific conditions.

In a previous study, toothbrush G improved the gingival condition of junior high school students who could not brush properly [17]. Our findings indicate that toothbrush G is less dependent on brushing technique. Since our results are consistent with the previous study, this evaluation method could be considered suitable for the functional evaluation of toothbrushes.

Few comprehensive and objective evaluation methods for toothbrush function have been reported using standardized evaluation methods. Evaluating toothbrush function using a single model is challenging due to varying oral cavity morphologies. However, our evaluation using multiple models, including concave, convex, and flat surfaces, makes it possible to cover most of the oral cavity morphologies. These experimental results were obtained using only two model types and therefore do not account for oral cavity morphology or the range of phenomena that occur during individual brushing motions. Future studies should determine whether findings from these models are reflected in the actual oral cavity.

## 5 Conclusion

This study examined a model developed for the objective evaluation of toothbrush function. In this study, functional evaluation was conducted using two models (smooth and interproximal surface, and SHC models) and two toothbrushes with different designs as test toothbrushes. To objectively evaluate the toothbrush, we measured penetration and cleaning efficiency at different angles, as well as the ability of the bristle tips to make contact with the SHC model surface. The use of these models to test various types of toothbrushes could facilitate the standardization of toothbrush functional evaluation, ultimately improving brushing techniques for effective oral hygiene.

## Data Availability

Raw data were generated at Nihon university, dentistry at Matsudo. Derived data supporting the findings of this study are available from the corresponding author M.Endoh on request.

## Acknowledgements

We would like to thank Editage (www.editage.jp) for English language editing.

## Author Contributions

〇Mami Endoh : Roles: Conceptualization, Data Curation, Investigation, Project administration, Resources, Supervision, Validation, Visualization, Writing – original draft –

〇Yuki Tsuji : Roles: Visualization, Writing – original draft –, Writing – review & editing

〇Tomoyo Jinushi : Roles: Investigation

〇Shota Kurihara: Roles: Investigation, Visualization

〇Takuro Higuchi : Roles: Methodology

〇Atsushi Takayanagi: Roles: Conceptualization, Formal analysis, Methodology, Validation, Writing – review & editing

〇Atsushi Yamagishi: Roles: Conceptualization, Investigation, Methodology, Validation, Visualization, Writing – review & editing

〇Takato Nomoto : Roles: Conceptualization, Funding acquisition, Project administration, Supervision, Writing – review & editing

## References

1. Cugini M, Warren PR. The Oral-B crossaction manual toothbrush: A 5-year literature review. JCDA. 2006;72:323.

2. Jansiriwattana W, Teparat-Burana T. Laboratory investigation comparing plaque removal efficacy of two novel-design toothbrushes with different brushing techniques. Dent J. 2018;6: 8. doi: 10.3390/dj6020008.

3. Otsuka R, Nomura Y, Okada A, Uematsu H, Nakano M, Hikiji K, et.al. Properties of manual toothbrush that influence on plaque removal of interproximal surface in vitro. J Dent Sci. 2020;15: 14–21.

4. Nakakura-Ohshima K, Hanasaki M, Nogami Y, Murakami N, Nakajima T, Hayasaki H. A novel method to evaluate interproximal accessibility of caregivers’ tooth brushing by using “invisibility” eyeglasses. Dent Oral Craniofac Res. 2018;4: 1–6. doi: 10.15761/DOCR.1000255.

5. Jinushi T, Endoh M, Shirata S, Yamagishi A, Takayanagi A, Nomoto T. A basic study on establishing on a toothbrush selection program for special needs patients, fourth report : force and conformity of toothbrushing. J Jpn soc disabil oral health. 2021;42: 235–242.

6. Endoh M, Jinushi T, Yamagishi A, Takayanagi A, Nomoto A. A study on the functional evaluation of toothbrushes: effects of toothbrush bristle hardness and brushing speed on cleaning results. Jpn J Dent Pract Adm. 2020;55: 61-68.

7. Endoh M, Jinushi T, Saegusa Y, Takayanagi A, Nomoto T. A basic study on the establishment of a toothbrush selection program for special needs patients, first report : cleaning ability and conformity of toothbrush bristles. J Jpn Soc Disabil Oral Health. 2020;41: 72-81.

8. Jinushi T, Endoh M, Takayanagi A, Saegusa Y, Inomata E, Ono A, et. al. A basic study to establish a toothbrush selection program for special needs patients: a second report-force and movement. J Jpn Soc Disabil Oral Health. 2021;42: 23-32.

9. Ono A, Endoh M, Jinushi T, Shirata S, Yamagishi A, Takayanagi A, et. al. A basic study on establishing a toothbrush selection program for special needs patients, fifth report: evaluating the cleaning ability by type of bristle ends and toothbrush size using an interproximal model. J Jpn Soc Disabil Oral Health. 2022;43: 90–100.

10. Endoh M, Jinushi T, Yamagishi A, Takayanagi A, Takato N. A functional evaluation of toothbrushes: effects of hardness and shape of toothbrush bristles on cleaning effect and tip movement. Jpn J Dent Pract Adm. 2021;55: 223–228.

11. Saegusa Y, Endoh M, Jinushi T, Shirata S, Yamagishi A, Takayanagi A, et al. A basic study on establishing on a toothbrush selection program for special needs patients third report: evaluating the cleaning ability among toothbrush sizes using a flat model. J Jpn Soc Disabil Oral Health. 2021;42: 160–169.

12. Endoh M, Ono A, Jinushi T, Shirata S, Yamagishi A, Takayanagi A, et. al. A functional evaluation of toothbrushes, third report: in vitro penetration performance used by interproximal model. Jpn J Dent Pract Adm. 2022;57: 39–46.

13. Kamijo Y. [Japanese permanent tooth anatomy]. Tokyo Anatome company. 1962. Japanese.

14. Takayanagi A. TDC post-graduate study seminar 2007. Necessary skills in and responsiveness of treatment in general dental practice. Preventive dentistry. Common sense and beyond in brushing: a scientific approach to successful brushing. Japanese.

15. Nakajima T, Nakakura-Ohshima K, Hanasaki M, Nogami Y, Murakami N, Nakamura Y, et.al. Difference of tooth brushing motion between dental hygienists and mothers-Focusing on self-tooth brushing and caregivers’-toothbrushing. Dent Oral Craniofac Res. 2017;4: 1–6.

16. Nakajima Y. Influences of brushing force and the number of brushing strokes on brushing effect during manual toothbrushing in adults. J Dent Health. 1971;21: 193–215.

17. Endoh M, Yamagishi A, Mikami N, Takayanagi A, Nomoto T. The effect of recommending toothbrushing for preventing gingivitis in oral health guidance, through cooperative school hours in a junior high school. Japanese J Dent Pract Adm. 2021;55: 217–222.

